# IDentif.AI: Artificial Intelligence Pinpoints Remdesivir in Combination with Ritonavir and Lopinavir as an Optimal Regimen Against Severe Acute Respiratory Syndrome Coronavirus 2 (SARS-CoV-2)

**DOI:** 10.1101/2020.05.04.20088104

**Authors:** Agata Blasiak, Jhin Jieh Lim, Shirley Gek Kheng Seah, Theodore Kee, Alexandria Remus, De Hoe Chye, Pui San Wong, Lissa Hooi, Anh T.L. Truong, Nguyen Le, Conrad E.Z. Chan, Rishi Desai, Xianting Ding, Brendon J. Hanson, Edward Kai-Hua Chow, Dean Ho

## Abstract

The emergence of severe acute respiratory syndrome coronavirus 2 (SARS-CoV-2) and coronavirus disease 2019 (COVID-19) has led to the rapid initiation of urgently needed clinical trials of repurposed drug combinations and monotherapies. These regimens were primarily relying on mechanism-of-action based selection of drugs, many of which have yielded positive in vitro but largely negative clinical outcomes. To overcome this challenge, we report the use of IDentif.AI, a platform that rapidly optimizes infectious disease (ID) combination therapy design using artificial intelligence (AI). In this study, IDentif.AI was implemented on a 12-drug candidate therapy search set representing over 530,000 possible drug combinations. IDentif.AI demonstrated that the optimal combination therapy against SARS-CoV-2 was comprised of remdesivir, ritonavir, and lopinavir, which mediated a 6.5-fold improvement in efficacy over remdesivir alone. Additionally, IDentif.AI showed hydroxychloroquine and azithromycin to be relatively ineffective. The identification of a clinically actionable optimal drug combination was completed within two weeks, with a 3-order of magnitude reduction in the number of tests typically needed. IDentif.AI analysis was also able to independently confirm clinical trial outcomes to date without requiring any data from these trials. The robustness of the IDentif.AI platform suggests that it may be applicable towards rapid development of optimal drug regimens to address current and future outbreaks.

## INTRODUCTION

Drug repurposing, or the use of approved and investigational therapies for other indications, has been a widely implemented strategy towards treating COVID-19. Examples include clinical studies of ritonavir and lopinavir (*1*); hydroxychloroquine in combination with azithromycin (*2*); favipiravir in combination with tocilizumab (NCT04310228); remdesivir (*3*); and losartan (NCT04312009), among others. In one trial, remdesivir met trial endpoints, reducing the median time to recovery from 15 days to 11 days (*P* < 0.001), and has ultimately received United States Food and Drug Administration (FDA) authorization for emergency use in severe COVID-19 patients (*4*). The majority of trial outcomes are either pending or have not shown clinical benefit over standard of care or placebo. As such, while drug repurposing enables rapid intervention against COVID-19, there is still a lack of clarity with regards to how to best treat this disease.

Traditional methods for implementing combination therapy and monotherapy based on drug repurposing rely on mechanism of action (MOA)-based drug selection and standard clinical dosing guidelines to achieve drug synergy and therapeutic efficacy. For example, a recent preclinical study showed that remdesivir as well as high-dose chloroquine were efficacious towards SARS-CoV-2 in vitro. While this is an established approach that has led to promising candidate therapies, many of these regimens were not able to translate their in vitro outcomes into successful clinical results. Therefore, optimal efficacy that is clinically relevant is a different objective that presents substantial challenges to traditional drug screening and repurposing methods. For example, if candidate effective drugs are given in combination at suboptimal respective doses, resulting efficacy is moderate or even absent. At the same time, the relative doses between drugs within a combination can substantially impact treatment efficacy and toxicity due to unpredictable drug interactions. Therefore, drug dosing has a critical role in identifying which drugs belong in the optimal combination in the first place. Therefore, optimizing treatment outcomes, particularly in combination therapy, ultimately relies on selecting the right drugs at the right respective doses (*5*, *6*). Reconciling drug-dose parameters also requires leveraging unpredictable drug interactions in order to mediate maximal efficacy of combination therapies. Unfortunately, simultaneously pinpointing these parameters is an extraordinarily complicated task. For example, a parameter space of 1 trillion (10^12^) possible combinations would be created from a pool of only 12 candidate therapies interrogated at 10 dose levels. This is an insurmountable barrier for traditional drug screening. Important studies have previously sought to leverage drug synergy interactions to predict multi-drug combinations (*7*). Other strategies have investigated higher order drug interactions to develop antimicrobial drug combinations (*8*). Bridging these findings with clinical validation remains a challenge due to the size of the experimental search space.

In this study, we sought to overcome these challenges in developing effective combination therapies against SARS-CoV-2 infection using the IDentif.AI platform. IDentif.AI harnesses a quadratic relationship between therapeutic inputs (e.g. drug and dose) and biological outputs (e.g. quantifiable measurements of efficacy, safety) to experimentally pinpoint optimal combinations from large parameter spaces with a marked reduction in the number of required experiments (Fig. 1). Identif.AI does not use pre-existing training datasets, but rather uses an orthogonally-designed set of calibrating regimens to simultaneously identify effective drugs and corresponding doses that optimize treatment outcomes from prohibitively large drug-dose parameter spaces that cannot be reconciled by brute force drug screening (*5*, *9*). In effect, IDentif.AI leverages these calibrating regimens to crowdsource SARS-CoV-2 live virus responses to experimentally drive the efficacy towards an optimal outcome. In this study, IDentif.AI was applied to a 12-drug set of candidate therapies to pinpoint clinically actionable combination therapy regimens against the live SARS-Cov-2 virus isolated from a nasopharyngeal swab of a patient in Singapore *(10)*. The 12-drug set included a broad spectrum of repurposed agents that are currently being evaluated in clinical studies for treatment of COVID-19 or being administered in conjunction with these therapies, including remdesivir (RDV), favipiravir (FPV), ritonavir (RTV), lopinavir (LPV), oseltamivir (OSV-P), dexamethasone (DEX), ribavirin (RBV), teicoplanin (TEC), losartan (LST), azithromycin (AZT), chloroquine (CQ), and hydroxychloroquine (HCQ). Based on prior studies of minimal resolution experimental design, 3 dosing levels were employed with these 12 drugs, creating a combinatorial space of 531,000 regimens (*11*). With a 3-order of magnitude reduction in required tests, we identified a clinically actionable list of 2-,3-, and 4-drug combinations ranked based on viral inhibition efficacy with accompanying safety data against kidney epithelial cells (Vero E6), liver epithelial cells (THLE-2) and cardiomyocytes (AC16). The top-ranked combination was comprised of remdesivir, ritonavir, and lopinavir, which mediated a 6.5-fold increase in efficacy (viral inhibition %) compared to remdesivir alone. Further demonstrating the clinical actionability of IDentif.AI, hydroxychloroquine and azithromycin combination was shown to be a relatively ineffective regimen, mirroring recent clinical results. Importantly, the IDentif.AI-pinpointed relative efficacy of the combinations and monotherapies was independently confirmatory of many of the clinical trial endpoints to date. These outcomes, coupled with the fact that foundational precursors to IDentif.AI have been clinically validated for infectious disease, oncology, and organ transplantation human studies, support the potential application of IDentif.AI as a clinical decision support platform for the optimized design of actionable combination therapy regimens *(12–14)*.

**Fig. 1.**
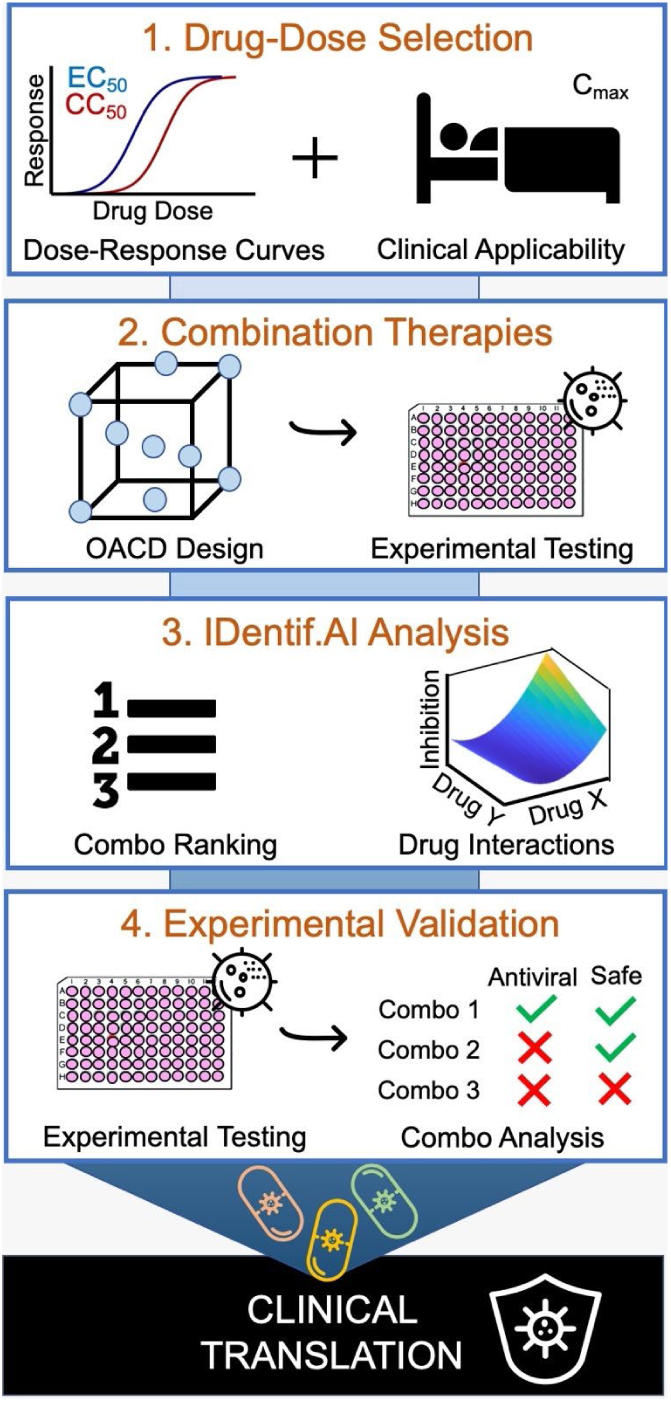
Project IDentif.AI workflow. Project IDentif.AI has four phases: (1) drug-dose selection established for each drug based on dose-response curves and C_max_ of clinically administered dosages, (2) combination therapies determined by a three-level orthogonal array composite design for experimental testing, (3) IDentif.AI analysis of the drug dose parameter space identifies drug-drug interactions and ranks optimal drug-dosage combinations, (4) experimental validation of IDentif.AI-designed and other selected combinations.

## MATERIALS AND METHODS

### IDentif.AI Analysis

IDentif.AI, a dynamic optimization AI-based platform, identifies the drug-dose parameter space by harnessing the quadratic relationship between biological responses to external perturbations, such as drug/dose inputs *(15)*. IDentif.AI analysis of the drug-dose parameter space identifies drug-drug interactions and ranks optimal drug-dosage combinations. This study aimed to use IDentif.AI to determine effective optimal drug-dosage combinations from a diverse set of 12 drugs currently being explored in clinical trials to combat the COVID-19 disease. The concentration levels of the 12 drugs for the in vitro IDentif.AI experiments were determined from EC_50_, CC_50_, and C_max_ for corresponding clinically administered dosages. From the in vitro experiment data, IDentif.AI analyses were performed to identify drug combinations from this pool of candidates that were effective against the SARS-CoV-2 virus.

Cell ATP activity for each well was processed using custom written Python 3.7.7 script (Python Software Foundation). Data were normalized to the average readout from the DMSO vehicle controls on the same plates. Vero E6, AC16 and THLE-2 %Cytotoxicity and viral activity %Inhibition (Vero E6) were calculated using the same formulae as for the drug monotherapy analysis (Supplementary Materials and Methods). %Inhibition calculations used cell and media only control wells. The resulting %Cytotoxicity and %Inhibition calculations were used as inputs in IDentif.AI analysis.

IDentif.AI analysis correlated drug combinations experimental results into a second-order quadratic series. Each independent drug combination inhibition and monotherapy inhibition replicate was used in the optimization process. The second-order quadratic model is as follows:

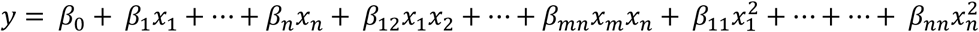

where *y* represents the desired biological response output (%Inhibition), *X_n_* is the *n*-th drug concentration, *β*_0_ is the intercept term, *β_n_* is the single-drug coefficient of the *n*-th drug, *β_mn_* is the interaction coefficient between the *m*-th and *n*-th drugs and *β_nn_* is the second-order coefficient for the *n*-th drug, while *m ≠ n*. This second-order quadratic analysis and parabolic response surface plot analysis were conducted using the built-in *“stepwiselm”* function in Matlab R2020a (Mathworks, Inc.) with custom-written code. IDentif.AI derived four quadratic series using bidirectional elimination approach with the *P* value from the F-Statistic as the selection criterion for the experimental results: %Inhibition, %Cytotoxicity, %Cytotoxicity AC16, and %Cytotoxicity THLE-2. Residual-based outlier analysis was performed for all four IDentif.AI series. Single replicates identified as outliers remained in the data set to account for biological variation. The combinations with all replicates identified as outliers were excluded from the data set and the IDentif.AI analysis was repeated.

IDentif.AI analysis yielded both drug-drug interaction plots and optimized drug combinations. The optimized drug combinations were ranked according to corresponding %Inhibition from the correlated second-order quadratic series with the %Cytotoxicity of the cell-lines (Vero E6, AC16, and THLE-2) serving as qualitative indicators for consideration. The predictive power was also calculated via adjusted R^2^ to establish the robustness of IDentif.AI optimization considering the number of drug and drug-drug interaction terms. Correlation coefficients were derived from the experimental output values and projected output values for the corresponding drug combinations.

### Statistical Analysis

All experiments were performed in at least triplicate biological repeats with data presented as means ± standard deviation (SD), unless otherwise stated. Shapiro-Wilk normality test was used to determine if samples were from normally distributed populations. Variance equality was tested with Bartlett’s test. The Kruskal-Wallis test by ranks was used for multiple comparisons, followed by Dunn’s post hoc test for pairwise comparisons. Student’s two-tailed t test and Wilcoxon rank sum test were used for comparing individual samples from normally and non-normally distributed populations, respectively. Bonferroni post-hoc correction was applied to account for multiple comparisons. Statistical analyses for coefficient estimation in the IDentif.AI analyses were performed using sum of squares F-test. Alongside the *P*-values, the results were interpreted in the light of logic, background knowledge and the specifics of the experimental design *(16)*.

## RESULTS

### Monotherapy Assessment and OACD Dataset Construction

A pool of drug candidates was first chosen and evaluated for downstream IDentif.AI analysis and drug combination optimization. The pool of candidate therapies for IDentif.AI-driven optimization contained eleven drugs that were hypothesized to inhibit SARS-CoV-2 viral infection *via* affecting: viral entry into the host cell - CQ, HCQ, AZT, LST, TEC; viral replication – RTV, LPV; viral RNA synthesis – RDV, FPV, RBV; viral release - OSV-P *(17–20)*. To create combinations actionable within the current clinical guidelines we aimed to investigate drug interaction space between the antiviral and concomitant medications. The last drug added to the pool, DEX, has been discussed for preventing and treating acute respiratory distress syndrome (ARDS) resulting from COVID-19 (*21*), NCT04325061. LST is a common hypertension drug whose dosing should not be paused while undergoing COVID-19 treatment (*22*). TEC is a wide spectrum antibiotic prescribed for pulmonary infections, potentially including those occurring as COVID-19-related complications *(23)*.

IDentif.AI interrogates drug-dose relationships in order to identify the most efficacious drug combinations within a defined drug concentration range. With the ultimate goal of clinical implementation, drug-dose response experiments were performed within concentration ranges that accounted for clinically implemented concentrations and avoided clinically unrealistic drug concentrations. The drugs with and without the addition of 100 TCID_50_ of SARS-CoV-2 virus were incubated with primate kidney cell line Vero E6 for 72h before measuring inhibition and cytotoxicity and generating the dose-response curves (Fig. S1).

Only high concentrations (>1 μM) of RDV, LPV, CQ and HCQ achieved half maximal absolute effective concentration (EC_50_) for the viral inhibition within the tested concentration ranges. High concentrations (>20 μM) of RTV, LPV and CQ led to half maximal absolute cytotoxic concentration (CC_50_) within the tested concentration ranges (Table 1). These results indicated low cellular effects of the selected monotherapies at the tested concentrations. No effect of the maximum vehicle concentration (0.1% DMSO) was detected on viral inhibition or on cytotoxicity (Student’s t-test, n = 12, *P* > 0.05). The EC_50_ and CC_50_ of HCQ, CQ, RDV, FPV, and RBV were different from previously reported values, attributable to differences in the experimental conditions (e.g. SARS-CoV-2 strain, assays, incubation periods) *(24*, 25). Because a common source of failure in translating in vitro results to clinical trials, including for coronaviruses, is the high ratio of EC_50_ to maximum plasma concentration (C_max_) achieved in the human body, C_max_ was included as a crucial consideration for selecting drug concentrations at Level 1 and Level 2 for each drug that ensure none of the drugs were overrepresented in relation to other drugs and to human pharmacokinetics (Table 1, Supplemental Results) (*26*). Regardless of the monotherapy antiviral activity, all drugs were considered for the combinatorial optimization process in order to identify possible unpredictable drug interactions that could markedly impact treatment efficacy and safety.

**Table 1.** Drug concentrations in combinatory treatment. Absolute half efficacy (EC_50_) and absolute half cytotoxicity (CC_50_) concentrations, and maximum plasma concentration (C_max_) and its source for each drug. NCT number is provided for COVID-19 clinical trials with drug dosages like those that where the basis for C_max_ selection. Concentration Levels 1 and 2 were based on: ^#^ absolute EC_10_ and absolute EC_20_ for RDV; ^*^ 2.5% and 5% of C_max_ for RTV, LST and TEC; and 5% and 10% of C_max_ for the rest of the drugs.

| Drug | $EC_{50}$<br>($\mu M$) | $CC_{50}$<br>($\mu M$) | $C_{max}$<br>($\mu M$) | COVID-19<br>Clinical Trial | Level 0<br>($\mu M$) | Level 1<br>($\mu M$) | Level 2<br>($\mu M$) |
| --- | --- | --- | --- | --- | --- | --- | --- |
| RDV | 1.1 | >100 | 9 (27) | NCT04292899 | 0 | 0.81# | 0.9 # |
| FPV | >600 | >600 | 331.83 (28) | NCT04310228 | 0 | 16.5915 | 33.183 |
| RTV | >100 | 97 | 20.39 (29) | - | 0 | 0.50975* | 1.0195* |
| LPV | 17 | 26 | 19.56 (30) | NCT04330690 | 0 | 0.978 | 1.956 |
| RBV | >100 | >100 | 17.3 (31) | NCT04276688 | 0 | 0.866 | 1.73 |
| CQ | 5.3 | 99 | 1.42 (32) | NCT04362332 | 0 | 0.071 | 0.142 |
| HCQ | 6.3 | >100 | 5.6 (33) | NCT04261517 | 0 | 0.28 | 0.56 |
| AZT | >100 | >100 | 0.32 (34) | NCT04329832 | 0 | 0.016 | 0.032 |
| OSV-P | >10 | >10 | 0.18 (35) | NCT04255017 | 0 | 0.009 | 0.018 |
| LST | >100 | >100 | 0.43 (36) | NCT04335123 | 0 | 0.01075* | 0.0215* |
| TEC | >50 | >50 | 20.475 (37) | - | 0 | 0.511875* | 1.02375* |
| DEX | >100 | >100 | 0.63 (38) | - | 0 | 0.0315 | 0.063 |

In order for IDentif.AI to determine optimized drug combinations from this 12-drug set, 100 drug-dose combinations were generated according to OACD (Table S1) and, together with drug monotherapies at concentration Level 1 and Level 2, were evaluated for their antiviral and cytotoxic activity on Vero E6 cells. Drug combinations’ cytotoxicity was additionally tested on human cell lines: liver (THLE-2), and cardiac myocytes (AC16). No effect of the maximum vehicle concentration (0.006% DMSO) was detected on viral inhibition or on cell cytotoxicity (Wilcoxon rank-sum test, n = 18, *P* > 0.05).

### IDentif.AI Analysis and Drug Combination Optimization

Utilizing the single drug and OACD drug treatment data, IDentif.AI analysis determined RDV/RTV/LPV to be the most efficacious 3-drug combination. It was also present in all top 10 ranked 4-drug combinations. RDV/LPV was the top ranked 2-drug combination (Table 2). While RDV was identified as the most efficacious single drug, in line with current clinical trial outcomes, IDentif.AI analysis determined that the 3-drug combination of RDV/RTV/LPV is critical for achieving maximal therapeutic efficacy. IDentif.AI analysis allows for comparative ranking of all possible combinations within the 12-drug set, including analysis of regimens currently being clinically investigated but that are not observed as top ranked optimized drug combinations. Both LPV/RTV (Kaletra®) and HCQ/AZT have been clinically evaluated as potential treatments against SARS-CoV-2 infection with discouraging outcomes. IDentif.AI analysis of these combinations revealed that they were identified to be sub-optimal - LPV/RTV ranked 1261 and HCQ/AZT ranked 5161 amongst all 9968 drug combinations that include up to 4-drugs, and predicted viral inhibition efficacies of 23% and 2%, respectively. The aforementioned findings were based on the IDentif.AI quadratic series assessing the %Inhibition experimental data with a close proximity as indicated by adjusted R^2^ of 0.898 (Table S2).

**Table 2.**
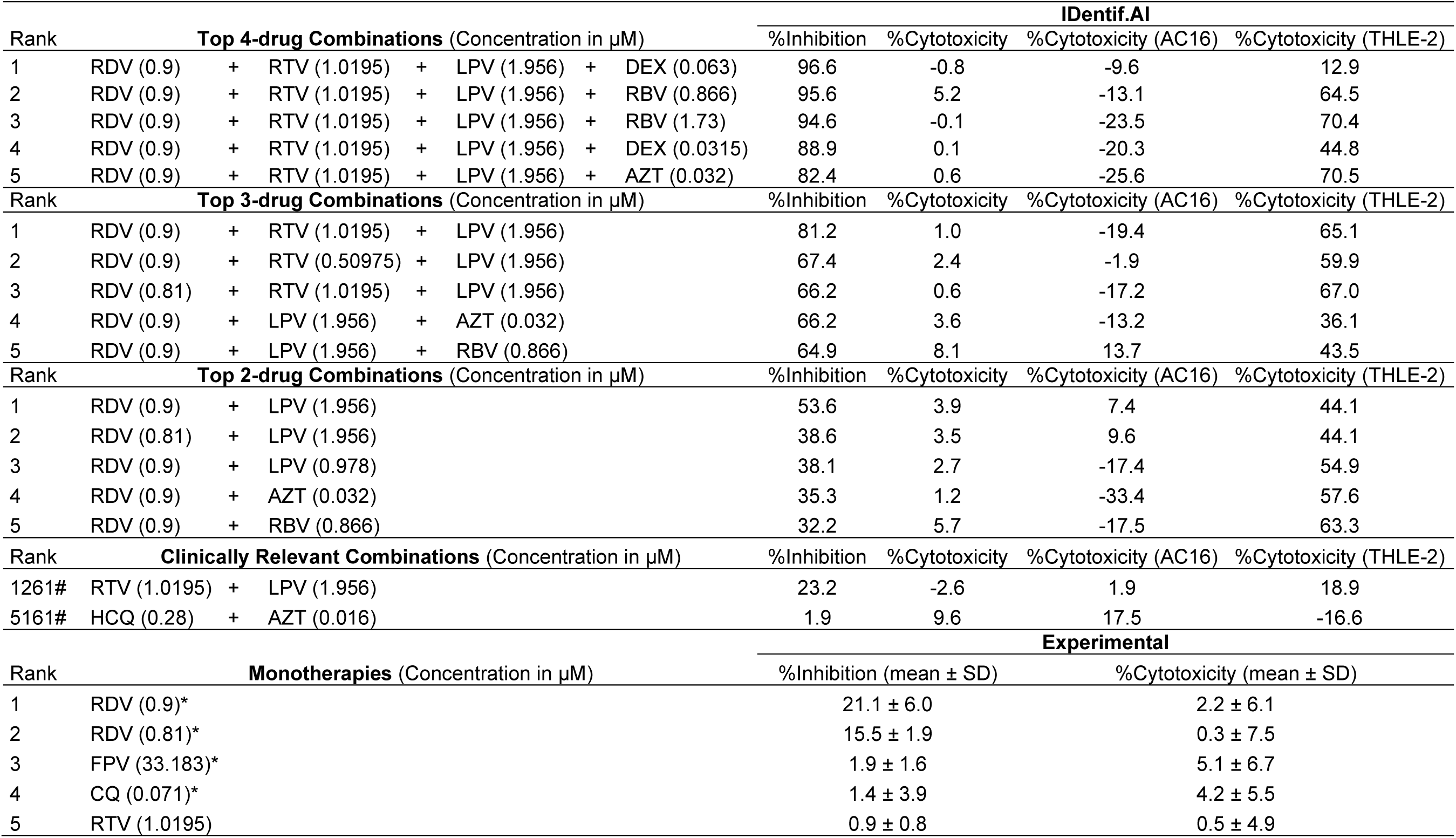
IDentfif.AI top ranked combinations. Top ranked IDentif.AI determined 4-drug, 3-drug, and 2-drug combinations with corresponding %Inhibition and %Cytotoxicity of Vero E6, AC16, and THLE-2. Monotherapies with corresponding %Inhibition and %Cytotoxicity Vero E6. Monotherapy experiments were run in triplicate. Data are shown as mean ± SD; *N=3*. # corresponds to rank of out 9968 combinations (4-drugs or less). * Indicates clinically tested monotherapies.

Multi-parameter IDentif.AI analysis allowed cytotoxicity of ranked combinations to be interrogated as well via deriving %Cytotoxicity quadratic series (Table S3-S5). The top ranked 3- and 4-drug RDV-based combinations were determined to have similar %Cytotoxicity in the Vero E6 cells in and experimentally measured single drug RDV treatments. IDentif.AI analysis also determined low %Cytotoxicity for the top 3-drug combination, RDV/RTV/LPV in the AC16 cells and higher %Cytotoxicity in the THLE-2 cells. This IDentif.AI-derived THLE-2 %Cytotoxicity was predicted to decrease with the addition of DEX in the top 4-drug combination (Table 2). Outlier analysis performed for each IDentif.AI quadratic series (Fig. S2-S5) identified and excluded OACD combination 15 from the AC16 %Cytotoxicity data set (Fig. S4) and combination 46 from the THLE-2 %Cytotoxicity data set (Fig. S5). These data sets were subsequently re-analyzed (Fig. S6-S7). Taken together, IDentif.AI identified RDV-based treatments as likely the most effective therapies against SARS-CoV-2 infections, with RDV/RTV/LPV capable of achieving maximal efficacy with potential reductions in overall toxicity if complemented with the fourth drug.

### Experimental Validation of IDentif.AI Results

Validation results were interpreted considering not only the *P*-values, but also the logic, background knowledge and specifics of the experimental design (*16*). The studies confirmed IDentif.AI ranking of RDV in combination with LPV and RTV as the optimal combination of the study, resulting in complete viral inhibition (Fig. 2A, Table S6). This combination resulted in a 6.5-fold increase in efficacy compared to RDV alone. RDV was confirmed as an essential driver of the antiviral efficacy in the optimized combinations, even though it mediated only moderate antiviral effect on its own. These validation studies were conducted following IDentif.AI identification of top-ranked optimized RDV-based drug combinations and comparative ranking of these combinations against other possible combinations.

**Fig. 2.**
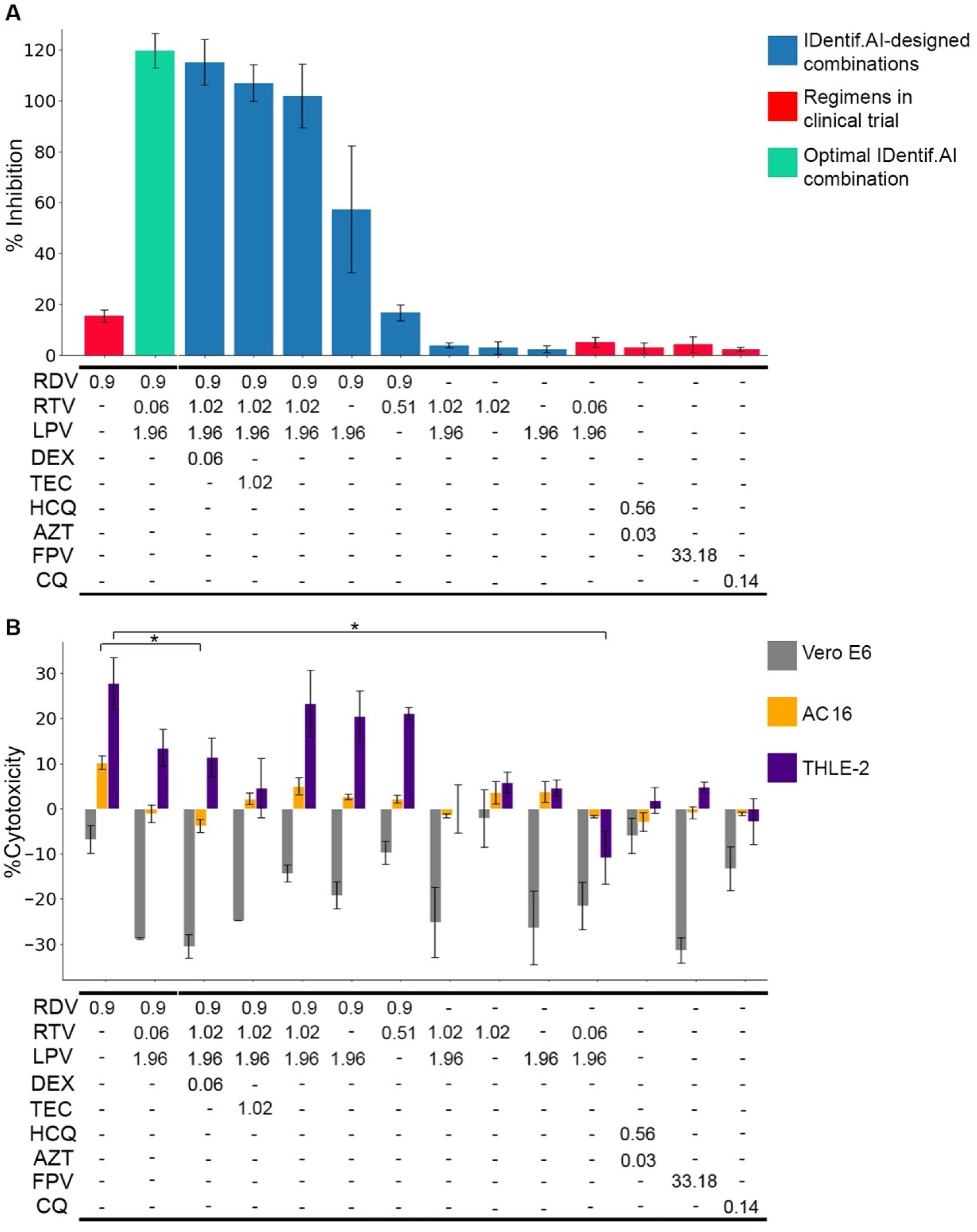
IDentif.AI-designed combinations validation. (**A**) %Inhibition of the optimal IDentif.AI combination of RDV/LPV/RTV (green), IDentif.AI designed combinations (blue), and regimens in clinical trials (red). (**B**) %Cytotoxicity of Vero E6 (grey), AC16 (orange), and THLE-2 (purple), of the optimal IDentif.AI combination of RDV/LPV/RTV, IDentif.AI designed combinations, and regimens in clinical trials. Data are shown as mean ± SD; *N=3. *P* < 0.05 (Kruskal Wallis test with Dunn’s post-hoc).

LPV and RTV are commonly administered together as RTV acts as a pharmacokinetic enhancer of LPV. Of note, the high antiviral effects of RDV/RTV/LPV were sustained when the RTV concentration was decreased 20-fold, and the RTV/LPV concentrations reflected the standard 100/400mg bid dosing in its clinically administered formulation (Kaletra®). As such, RDV/RTV/LPV combination likely does not require increasing RTV dose beyond what is commonly used clinically and is readily clinically actionable upon an approval. Importantly, RTV/LPV synergistic effects were limited to antiviral inhibition and did not adversely affect RDV cytotoxicity. RDV’s cytotoxicity, also reported clinically (*27*), was not enhanced in any of the combinations (Fig. 2B). In fact, the results suggest that RDV/RTV/LPV may suppress RDV-induced cytotoxicity both on human cardiac myocytes and human liver cell lines, with this effect being strengthened when combining with concomitant drugs, DEX and TEC. Monkey kidney cells did not exhibit cytotoxicity after any of the treatments. While these data suggest that specific top-ranked RDV-based combinations may also lower RDV-induced cytotoxicity, this observation requires further investigation. Dose limiting and drug exclusion experiments further deciphered the contribution of each drug towards overall RDV/RTV/LPV antiviral activity. RDV was confirmed to have the greatest contribution, with LPV/RTV on its own not mediating viral inhibition. High concentrations of LPV were critical to maximizing the RDV/RTV/LPV antiviral activity. While the concentration of RTV was not a critical determinant of the resulting efficacy of the combination, the presence RTV was critical to RDV/RTV/LPV achieving maximal viral inhibition. Further confirming the accuracy of IDentif.AI analysis, validation of currently clinically trialed treatments against COVID-19, LPV/RTV (Kaletra®)(1), HCQ/AZT *(39, 40)*, FPV (NCT04310228) and CQ (NCT04362332) did not induce as much viral inhibition as compared to RDV alone. These data confirm that IDentif.AI can accurately reflect the unsatisfactory outcomes observed in those clinical trials, without incorporating any prior clinical data or drug mechanism assumptions as input.

Drug-drug interaction analysis of IDentif.AI results were also compared to experimental observations and known clinical investigations. %Inhibition IDentif.AI response surface plot mirrored well-documented and experimentally confirmed synergy between RTV and LPV (Fig. 3A). In contrast, IDentif.AI identified an antagonistic interaction between RTV and OSV-P (Fig. 3B), a combination that is currently being investigated in clinical trials (NCT04303299). Combining RDV with LPV only, which to our knowledge has not been explored clinically as a registered trial, doubled their individual viral inhibition when added together. Accordingly, the corresponding %Inhibition IDentif.AI response surface plot identified a previously unknown synergistic interaction between RDV and LPV (Fig. 3C). Further confirming IDentif.AI rankings and validation experiments, the RDV/RTV interaction was not significant, but when given in 3-drug combination, RTV boosted the RDV/LPV interaction almost two times (Fig. 3D). These results further highlight the ability of IDentif.AI to leverage unexpected drug-dose interactions to identify optimal drug combinations from a massive drug-dose search space.

**Fig. 3.**
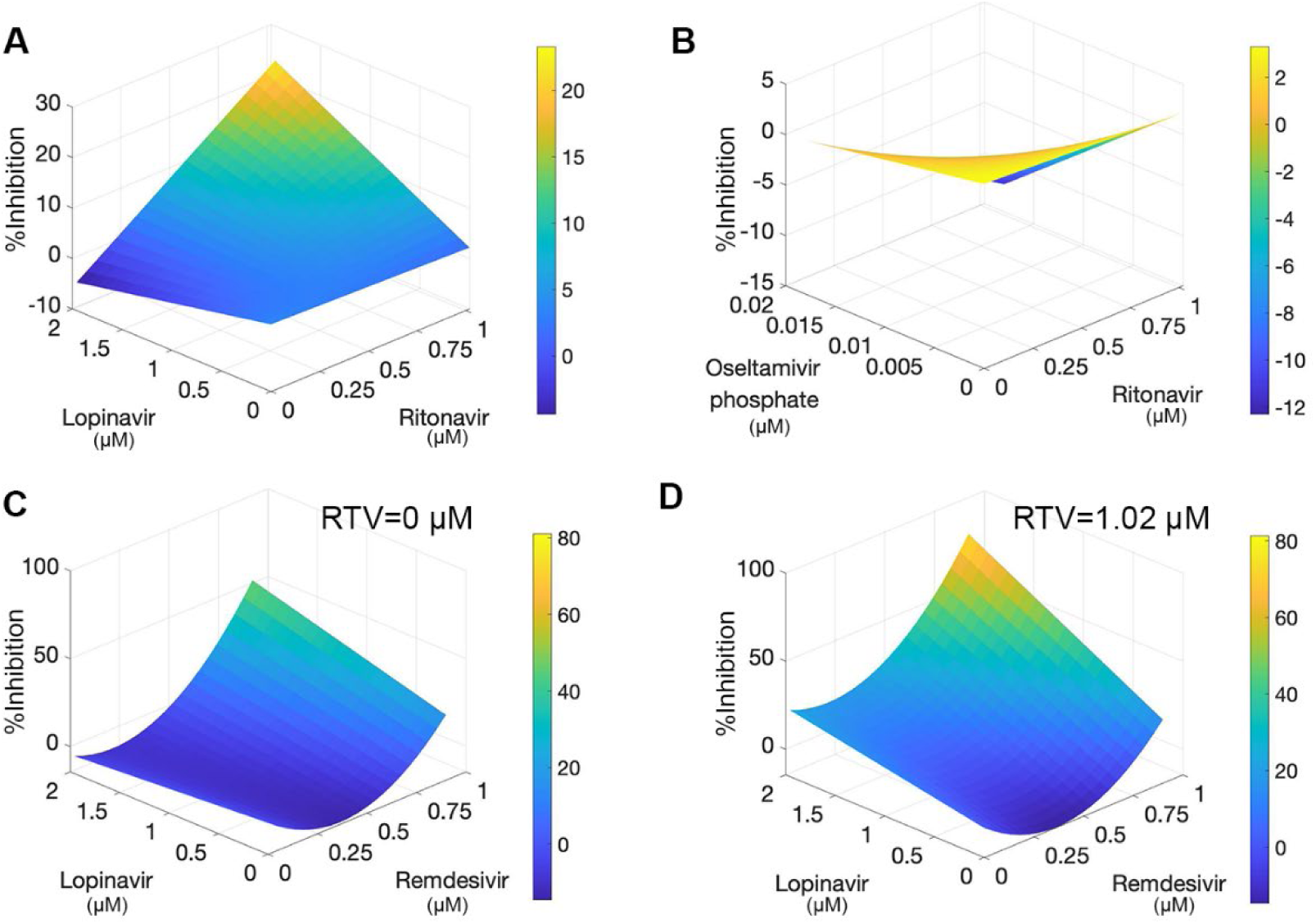
Antiviral drug interactions determined from the IDentif.AI analysis based on OACD concentration Levels 1 and 2. (**A**) IDentif.AI determined synergistic interaction between lopinavir and ritonavir. (**B**) Antagonistic interaction between ritonavir and oseltamivir phosphate. (**C-D**) Synergistic interaction between remdesivir and lopinavir (C) was boosted by the presence of ritonavir (RTV) (D).

## DISCUSSION

This study harnessed the IDentif.AI platform to interrogate a 12 drug-dose parameter space against the SARS-CoV-2 live virus to develop actionable and optimized combination therapy regimens. IDentif.AI implementation addresses several important factors when designing multi-drug regimens that are best suited for clinical translation from in vitro validation, especially under urgent scenarios like COVID-19. Importantly, IDentif.AI considers the critical need for simultaneous reconciliation of drug composition and dosing within combination therapy design. MOA-based drug selection alone followed by dose finding, while an established method of combination therapy design, presents substantial barriers to the optimization process since drug dosing also plays a role in determining which drugs belong in an ideal combination. In lieu of validating a small number of MOA-based potential drug combinations for efficacy which is commonly observed in traditional workflows, IDentif.AI takes an MOA-agnostic approach to efficiently analyze crowdsourced therapeutic responses to the live virus following expansive drug-dose exposure to both outline the drug-dose space and define resulting drug-dose compositions of the optimal regimens (*41*). With this data, IDentif.AI is able to leverage on unexpected dose-dependent drug interactions to mediate improved treatment outcomes over MOA-based drug selection followed by dose finding.

Another critical aspect of IDentif.AI is that the in vitro drug dosing parameter space interrogated in this study is a departure from traditional drug screening approaches. In traditional drug screening, compounds that do not elicit at least a low micromolar EC_50_ treatment response during drug dose-response evaluations are typically removed from further consideration, thereby markedly reducing the number of candidate therapies and possible drug combinations. The removal of these drug candidates is a key driver of sub-optimal treatment responses as it ignores a broad spectrum of potential combinations that can be assessed. Lack of monotherapy efficacy does not preclude the use of these drug candidates from IDentif.AI’s combinatorial search space. Instead, IDentif.AI’s approach allows for continued evaluation of these drugs to determine if they are vital toward driving previously unknown drug interactions that optimize combinatorial treatment outcomes. In addition to being observed in this study, this phenomenon has also been observed with our prior clinical studies in chronic infectious diseases and blood and solid cancers, among other indications (*5*, *8, 9, 42)*.

The outcome of applying IDentif.AI towards combating SARS-CoV-2 infection is an extensive list of combinations ranked by efficacy and/or safety that can be queried by a clinician based on clinically actionable criteria. These include, but are not limited to: highest ranked 2-, 3-, 4-drug combinations by efficacy; highest ranked combinations that do not contain certain drugs due to supply shortages; highest ranked combinations that do not contain certain drugs or contain lower dosages of certain drugs due to patient co-morbidities; and highest ranked combination comprised of only approved therapies, among others. In the context of optimized regimen design, which assesses regimen performance from the entire landscape of possible drug/dose parameters, IDentif.AI-enabled comparative evaluation of the relative efficacy of a broad spectrum of optimized regimens and clinically investigated regimens also independently confirmed the reported outcomes of clinical trials. This provides additional support for the potential application of IDentif.AI as a clinical decision support platform. For example, the relatively low efficacy exhibited HCQ alone (3.9%) or by HCQ and AZT (3% inhibition) in this study aligned with recent reporting of clinical outcomes for this drug given in mono- and combinatory therapy (*39*, *40)*. The relatively low efficacy (3.9% and 5.2%) of RTV and LPV combination when assessed by IDentif.AI at two different dosing ratios also aligned with recently reported outcomes showing no benefit over standard care (*1*). IDentif.AI also revealed a relatively low efficacy of FPV monotherapy (1.9% inhibition) and various combinations. This was consistent with clinical findings of FPV being potentially clinically effective only when administered with interferon-alpha, not included within our drug library (*43*). Of note, RDV alone resulted in the highest relative efficacy for monotherapy (15.5%) in this study. To date, compassionate use of RDV resulted in clinical improvement of 68% of the patients, and a larger study resulted in a statistically significant improvement in median time to recovery from 15 days to 11 days (*4*). At the same time, an RDV study in severe COVID-19 patients was also recently terminated early (NCT04257656). Nonetheless, it has received FDA authorization for emergency use in severe COVID-19 patients. The substantial difference in efficacy observed between sub-optimal and optimal regimens highlights the importance of leveraging platforms such as IDentif.AI to systematically design combination therapies. This capability, along with the potentially predictive capacity of IDentif.AI for clinical trial outcomes could provide clinicians with an expanded arsenal of evidence-based candidate treatments and important insights into which potential treatments to further evaluate or potentially avoid under time-sensitive circumstances.

It is important to note that the results reported here are derived from primarily an in vitro SARS-CoV-2 study. Further clinical validation of the outlined combinations in randomized controlled trials will be needed. It should also be noted that, while RDV did not mediate a significant clinical benefit in severe COVID-19 patients, its efficacy in patients with varying disease burden severities should be evaluated further. Furthermore, the mixed reported clinical outcomes support the need for improved regimen design of RDV-based treatment. In the event of downstream clinical validation of IDentif.AI-designed combinations, the drug dosage ratios within the combination may vary from those pinpointed by IDentif.AI. In addition, it is possible that the optimal drug combinations may vary between patients due to their severity of infection, comorbidities, and other factors. It is for these reasons that potential downstream trials may be effective at determining the potential clinical benefit of the IDentif.AI-designed combinations if the enrolled patients are stratified by these aforementioned clinical parameters. The 12-drug search set used in this study did not include every therapeutic option currently under clinical investigation. Additional studies, including other repurposed compounds, may yield additional highly ranked and effective combination regimens. Also, as IDentif.AI can be applied to novel small molecules and antibody therapies, their inclusion into the drug pool would add further insight into other potentially actionable regimens. Furthermore, given the rapid mutagenicity of RNA viruses like SARS-CoV-2, future studies with different drug candidates and different SARS-CoV-2 strains may yield different combinations. However, the efficiency and deterministic nature of IDentif.AI allows it to derive a ranked list of optimal regimens from a given set of drug candidates against a defined in vitro infectious disease model within two weeks. This further supports its potential application as a clinical decision support platform for the optimized design of combination therapy regimens against multiple SARS-CoV-2 strains as well as future unknown pathogens that will again require rapid mobilization and clinical guidance for effective treatment options.

## Data Availability

All relevant data has been included in the supplementary section. The authors can be contacted if additional information is needed.

## ACKNOWLEDGMENTS

D.H. gratefully acknowledges support from the Office of the President, Office of the Senior Deputy President and Provost, and Office of the Deputy President for Research and Technology at the National University of Singapore. D.H. also gratefully acknowledges the Ministry of Education Tier 1 FRC Grant. D.H. and E.K.C. gratefully acknowledge the National Research Foundation Singapore under its AI Singapore Programme (Award Number: *AISG-GC-2019-002)*, and Singapore Ministry of Health’s National Medical Research Council under its Open Fund-Large Collaborative Grant (“OF-LCG”)(MOH-OFLCG18May-0028). E.K.C. is supported by the National Research Foundation Singapore and the Singapore Ministry of Education under its Research Centres of Excellence Initiative (Cancer Science Institute of Singapore RCE Main Grant), Ministry of Education Academic Research Fund (MOE AcRF Tier 2 [MOE2019-T2-1-115]), Singapore Ministry of Health’s National Medical Research Council under its Open Fund-Large Collaborative Grant (“OF-LCG”)(MOH-OFLCG18May-0023 and MOH-OFLCG18May-0028) and National Research Foundation Competitive Proton Research Programme (NRF-CRP-2017-05). X.D. acknowledges support from National Key Research and Development Program of China (2017ZX10203205) and National Science Foundation of China (81871448). S.G.K.S., D.H.C., P.S.W., C.E.Z.C. and B.J.H gratefully acknowledge funding support from Future Systems and Technology Directorate, Singapore Ministry of Defence.

## SUPPLEMENTARY MATERIALS

Supplementary Materials and Methods

Supplementary Results

Figs. S1-S7

Tables S1-S6

References 1-23

